# Evolution and epidemic spread of SARS-CoV-2 in Brazil

**DOI:** 10.1101/2020.06.11.20128249

**Authors:** Darlan S. Candido, Ingra M. Claro, Jaqueline G. de Jesus, William M. Souza, Filipe R. R. Moreira, Simon Dellicour, Thomas A. Mellan, Louis du Plessis, Rafael H. M. Pereira, Flavia C. S. Sales, Erika R. Manuli, Julien Thézé, Luiz Almeida, Mariane T. Menezes, Carolina M. Voloch, Marcilio J. Fumagalli, Thais M. Coletti, Camila A. M. Silva, Mariana S. Ramundo, Mariene R. Amorim, Henrique Hoeltgebaum, Swapnil Mishra, Mandev S. Gill, Luiz M. Carvalho, Lewis F. Buss, Carlos A. Prete, Jordan Ashworth, Helder Nakaya, Pedro S. Peixoto, Oliver J. Brady, Samuel M. Nicholls, Amilcar Tanuri, Átila D. Rossi, Carlos K.V. Braga, Alexandra L. Gerber, Ana Paula Guimarães, Nelson Gaburo, Cecila S. Alencar, Alessandro C.S. Ferreira, Cristiano X. Lima, José Eduardo Levi, Celso Granato, Giula M. Ferreira, Ronaldo S. Francisco, Fabiana Granja, Marcia T. Garcia, Maria Luiza Moretti, Mauricio W. Perroud, Terezinha M. P. P. Castineiras, Carolina S. Lazari, Sarah C. Hill, Andreza A. de Souza Santos, Camila L. Simeoni, Julia Forato, Andrei C. Sposito, Angelica Z. Schreiber, Magnun N. N. Santos, Camila Zolini de Sá, Renan P. Souza, Luciana C. Resende-Moreira, Mauro M. Teixeira, Josy Hubner, Patricia A. F. Leme, Rennan G Moreira, Maurício Lacerda Nogueira, CADDE-Genomic-Network, Neil M Ferguson, Silvia F. Costa, José Luiz Proenca-Modena, Ana Tereza R. Vasconcelos, Samir Bhatt, Philippe Lemey, Chieh-Hsi Wu, Andrew Rambaut, Nick J. Loman, Renato S. Aguiar, Oliver G. Pybus, Ester C. Sabino, Nuno Rodrigues Faria

**Author notes:** These authors contributed equally to this work (Darlan S. Candido, Ingra M. Claro, Jaqueline G. de Jesus, William M. Souza, Filipe R. R. Moreira, Simon Dellicour, Thomas A. Mellan).

## Abstract

Brazil currently has one of the fastest growing SARS-CoV-2 epidemics in the world. Due to limited available data, assessments of the impact of non-pharmaceutical interventions (NPIs) on virus transmission and epidemic spread remain challenging. We investigate the impact of NPIs in Brazil using epidemiological, mobility and genomic data. Mobility-driven transmission models for São Paulo and Rio de Janeiro cities show that the reproduction number (*R*_*t*_) reached below 1 following NPIs but slowly increased to values between 1 to 1.3 (1.0–1.6). Genome sequencing of 427 new genomes and analysis of a geographically representative genomic dataset from 21 of the 27 Brazilian states identified >100 international introductions of SARS-CoV-2 in Brazil. We estimate that three clades introduced from Europe emerged between 22 and 27 February 2020, and were already well-established before the implementation of NPIs and travel bans. During this first phase of the epidemic establishment of SARS-CoV-2 in Brazil, we find that the virus spread mostly locally and within-state borders. Despite sharp decreases in national air travel during this period, we detected a 25% increase in the average distance travelled by air passengers during this time period. This coincided with the spread of SARS-CoV-2 from large urban centers to the rest of the country. In conclusion, our results shed light on the role of large and highly connected populated centres in the rapid ignition and establishment of SARS-CoV-2, and provide evidence that current interventions remain insufficient to keep virus transmission under control in Brazil.

**One Sentence Summary:** Joint analysis of genomic, mobility and epidemiological novel data provide unique insight into the spread and transmission of the rapidly evolving epidemic of SARS-CoV-2 in Brazil.

## Main Text

Severe acute respiratory syndrome coronavirus type 2 (SARS-CoV-2) is a novel zoonotic betacoronavirus with a 30-kb genome that was first reported in December 2019 in Wuhan, China (*1, 2*). As of 9 June 2020, coronavirus disease (COVID-19) has caused over 7 million cases and 404 thousand deaths globally (*3*). SARS-CoV-2 was declared a public health emergency of international concern on 30 January 2020. The virus can be classified into two phylogenetic lineages, named lineage A and B, that spread from Wuhan before strict travel restrictions were enacted (*4, 5*) and now co-circulate around the world (*6*). The case fatality rate of SARS-CoV-2 infection has been estimated between 1.2 and 1.6% (*7-9*) with substantially higher ratios in those aged above 60 years (*10*). Over 18 to 56% of SARS-CoV-2 transmission is from asymptomatic or pre-symptomatic individuals (*11-13*), complicating epidemiological assessments and public health efforts to curb the pandemic.

While the growth of the SARS-CoV-2 epidemics in China, Italy, and Spain is declining in response to NPIs (*3*), the number of SARS-CoV-2 cases and deaths in Brazil continues to increase (*14*) (**Fig. 1A**). Brazil has now reported 739,503 SARS-CoV-2 cases, the second largest number in the world, and 38,406 deaths (as of 9 June 2020). Over one third of the cases (35%) in Brazil are concentrated in southeast region which includes São Paulo city (**Fig. 1B**), the world’s fourth largest conurbation, where the first case in Latin America was reported on 25 February 2020 (*15*). Diagnostic assays for SARS-CoV-2 molecular detection were widely distributed across the national public health laboratory network from early on (*16, 17*). However, several factors including delays in reporting, changes in notification and heterogeneous access to testing across populations, have obfuscated real-time assessments of virus transmissibility based on SARS-CoV-2 case counts (*15*). Given this, death counts caused by severe acute respiratory infections (SARI) reported to the Sistema Único de Saúde (SUS) provide a more accurate measure for SARS-CoV-2 transmission in the country (*18*). Changes in SARS-CoV-2 transmissibility are strongly associated with changes in average mobility (*19, 20*), and can typically be measured by calculating the time-varying reproduction number, *R*_*t*_, defined as the average number of secondary infections caused by an infected person. *R*_*t*_ >1 indicates a growing epidemic while *R*_*t*_ <1 is needed to achieve a decrease in transmission.

**Fig. 1.**
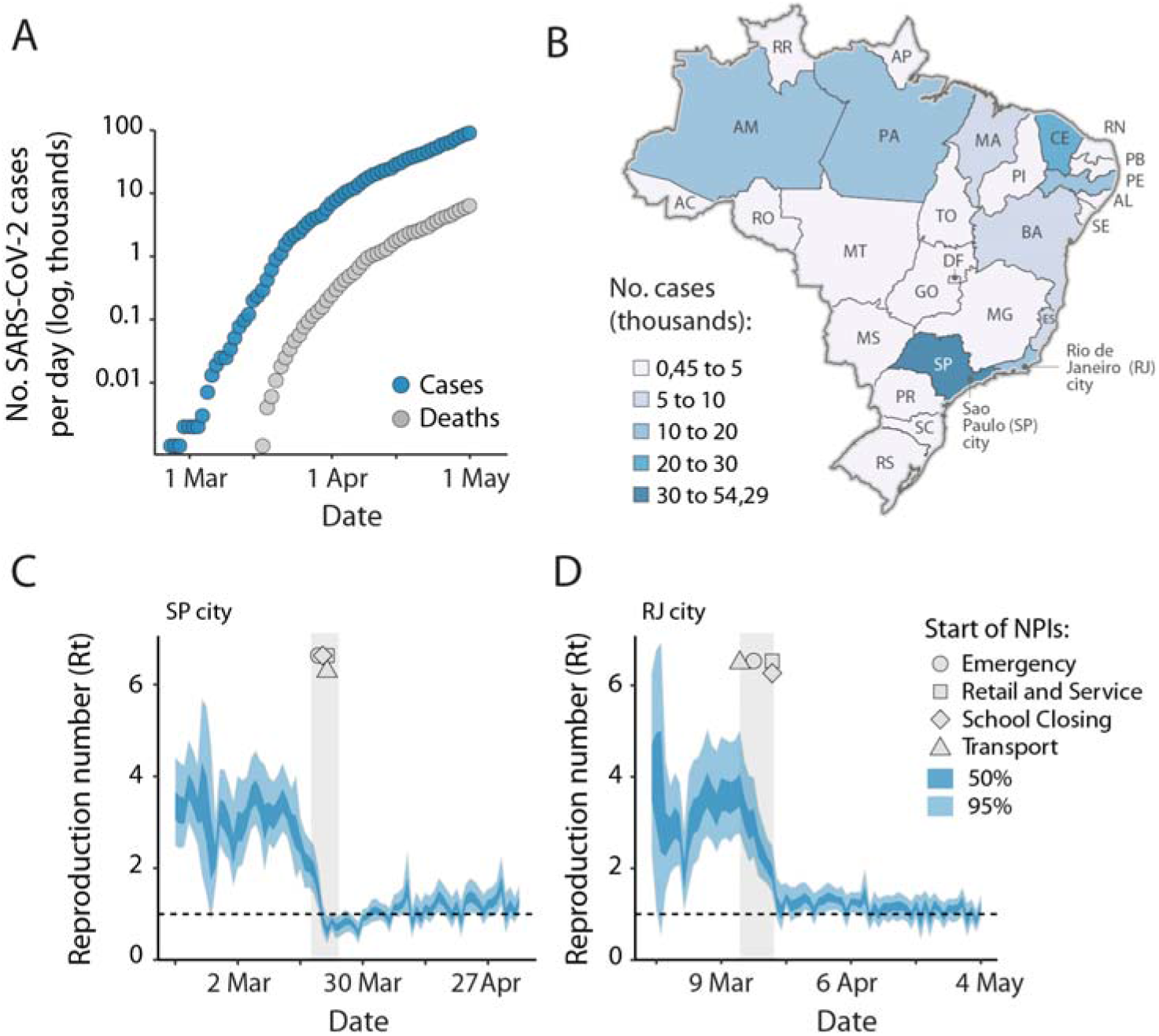
SARS-CoV-2 epidemiology and epidemic spread in Brazil. **(A)** Cumulative number of SARS-CoV-2 reported cases (blue) and deaths (grey) in Brazil. **(B)** States are coloured according to the number of cumulative confirmed cases by April 30, 2020. (**C** and **D**) Reproduction number *Rt* over time for the cities of São Paulo (**C**) and Rio de Janeiro (**D**). Reproduction numbers (*R*_*t*_) were estimated using a Bayesian approach incorporating daily number of deaths and four variables related to mobility data (a social isolation index from Brazilian geolocation company *InLoco*, and Google mobility indices for time spent in transit stations, parks, and the average between groceries and pharmacies, retail and recreational, and workspaces). Dashed horizontal line indicates *Rt*=1. Grey area and geometric symbols represent the time at which NPIs interventions were implemented. Bayesian credible intervals (BICs, 50 and 95%) are shown as shaded areas. The 2-letter ISO 3166-1 codes for the 27 federal units in Brazil (26 federal states and 1 federal district) are as follows: AC=Acre, AL=Alagoas, AM=Amazonas, AP=Amapá, BA=Bahia, CE=Ceará, ES=Espírito Santo, DF=Distrito Federal, GO=Goiás, MA=Maranhão, MG=Minas Gerais, MS=Mato Grosso do Sul, MT=Mato Grosso, PA=Pará, PB=Paraíba, PE=Pernambuco, PI=Piauí, PR=Paraná, RJ=Rio de Janeiro, RN=Rio Grande do Norte, RO=Rondônia, RR=Roraima, RS=Rio Grande do Sul, SC=Santa Catarina, SE=Sergipe, SP=São Paulo, TO=Tocantins.

We used a Bayesian semi-mechanistic model (*21, 22*) based on SARI deaths and human mobility data to investigate daily changes in *R*_*t*_ in São Paulo city (12,2 millions inhabitants) and Rio de Janeiro city (6,7 millions inhabitants), the largest urban metropoles in Brazil (**Figs. 1C** and **1D**). Non-pharmaceutical policies in Brazil consisted of school closures implemented between 12 and 23 March 2020 across the countries’ 27 federal units/states, and stores closures implemented between 13 and 23 March 2020. In São Paulo city, schools started closing on the 16 March and stores closed four days later. At the start of the epidemics, we found *R*_*t*_ *>3* in São Paulo and Rio de Janeiro, and that concurrent with the timing of state mandated non-pharmaceutical interventions (NPIs), *R*_*t*_ values fall close to 1.

Analysis of the reproduction number after NPI implementation highlights several notable mobility-driven features. We find that *R*_*t*_ drops during weekends and holidays due to differences in movement patterns and social isolation. For example, after the implementation of NPIs in São Paulo, the mean *Rt* was 1.0 (95% Bayesian credible interval: 0.7–1.3) during weekends, compared to 1.1 (95% BCI: 0.9–1.5) during weekdays. In addition, there was a period immediately following NPIs, between 21 March and 31 March 2020, where the *R*_*t*_ was consistently less than 1 in São Paulo (**Fig. 1C**). However, after this initial drop, the *R*_*t*_ in São Paulo rises above 1 and shows an increasing trend associated with increased population mobility. For example, this can be seen in the Google transit stations index, that rises from -60% to -52%, and by the decrease in the social isolation index from 54% to 47%. By 4 May 2020, the reproduction number had an estimated value of 1.3 (CI95%: 1.0, 1.6) in both São Paulo and Rio de Janeiro cities (**Table S1**). However, we point out there were instances in the previous 7 days where the 95% credible intervals for *R*_*t*_ had fallen below a value of 1, drawing attention to the fluctuations and uncertainty in the reproduction number for both cities.

Early sharing of genomic sequences, including the earliest released SARS-CoV-2 genome, Wuhan-Hu-1, on 10 January (*23*), has enabled unprecedented global preparedness and response (*24, 25*). However, despite the thousands of virus genomes deposited on public access databases, there is a lack of consistent sampling structure that hampers accurate reconstructions of virus movement and transmission using phylogenetic analyses. To investigate how SARS-CoV-2 became established across Brazil, and to quantify the impact of NPIs in virus spatiotemporal spread, we tested a total of 26,732 samples from public and private laboratories using real-time PCR assays and found 7,944 (29%) to be positive for SARS-CoV-2. We then focused our sequencing efforts on generating a spatially representative genomic dataset with curated metadata that maximized the association between the number of sequences and the number of SARS-CoV-2 confirmed cases per state.

We generated 427 SARS-CoV-2 new genomes from Brazil with >75% 200-fold genome coverage that were sampled between the 5 March to 30 April 2020 (**Figs. S1** and **S2** and **Data S1**). The data generated here originated from 84 municipalities across 18 of 27 federal units spanning all regions in Brazil (**Fig. 2A, Fig. S2**). Sequenced samples were obtained from samples collected on average 5 days (median, range: 0 to 29 days) after onset of symptoms and were generated in 3 laboratories using harmonized sequencing and bioinformatic protocols. Despite the use of different diagnostic assays and sequencing primers across sites (**Table S2)**, our data suggests that higher genome coverage samples are characterized by lower real-time PCR cycle threshold (Ct) and shorter times between symptom onset and sample collection (**Fig. S3**). When we include 63 additional available sequences from Brazil deposited in GISAID, a strong spatial representativity was obtained when considering the total number of genomes per state and SARI SARS-CoV-2 cases (*n*=490 sequences from 21 states, Spearman’s correlation, ρ=0.83, *P*-value < 3.79 ⨯ 10^×7^, **Fig. 2A**). This correlation ranged from 0.70 to 0.83 (*P*-values < 0.0001) when considering SARS-CoV-2 deaths or suspected cases, respectively (**Fig. S4**). Moreover, we used an *in-silico* assessment of diagnostic assay specificity for Brazilian strains (*n=*490) to identify potential mismatches in some assays targeting Brazilian strains. We find that the forward primers of the Chinese CDC and Hong Kong University nucleoprotein-targeting RT-qPCR may be less appropriate for use in Brazil than other diagnostic assays for which few or no mismatches were identified (**Fig. S5, Table S3**).

**Fig. 2.**
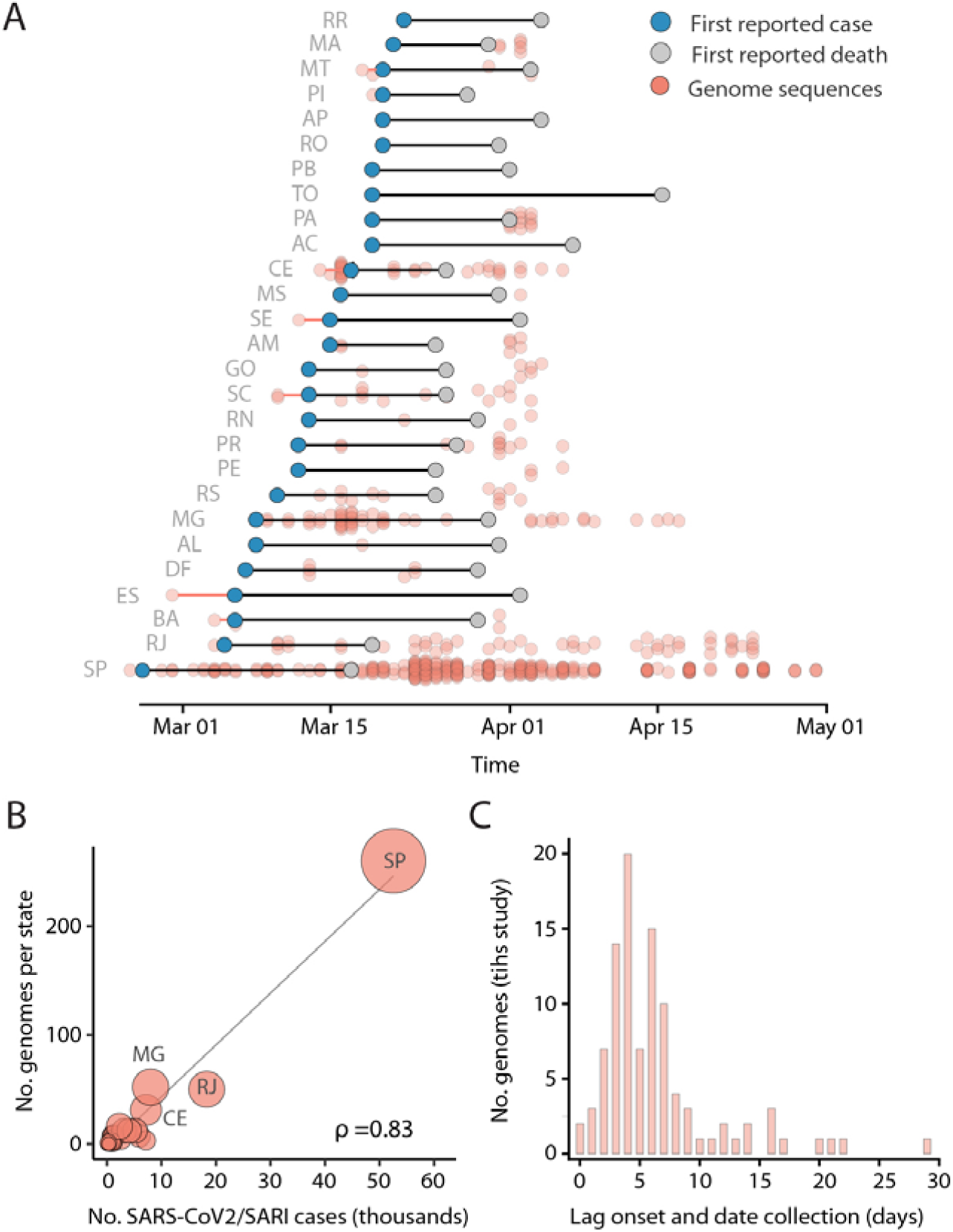
Spatial representativity in genomic sampling. **(A)** Dumbbell plot showing temporal intervals between date of collection of sampled genomes, notification of first cases and first deaths in each state. Red lines indicate lag between date collection of first genome sequence and first reported case. The 2-letter ISO 3166-1 codes for the 27 federal units in Brazil (26 federal states and 1 federal district) are shown in the caption of Fig. 1. **(B)** Spearman’s rank (ρ) correlation between the number of SARS-CoV-2 confirmed and suspected SARI cases against number of sequences for each of the 21 Brazilian states included in this study (see also Fig. S2). Circle sizes are proportional to the number of sequences for each federal unit. (**C**) Interval between the date of symptom onset and the date of sample collection for the sequences generated in this study.

The time between the date of the first reported case and the date of collection of the first sequence analysed for each state was 4.5 days (**Fig. 2A**). For 8 federal states genomes were obtained from samples collected up to 6 days before the first case notifications. We found that most (*n*=485/490) Brazilian sequences belong to SARS-CoV-2 lineage B, with only 5 strains belonging to the lineage A (2 from Amazonas, 1 from Rio Grande do Sul, 1 from Minas Gerais and 1 from Rio de Janeiro) (**Data S1 and S2** and **Fig. S6** shows detailed lineage information for each sequence).

We used a combination of genomic, spatial, mobility and epidemiological data to investigate changes in transmission patterns in Brazil over time. We first estimated the temporal signal of our global alignment by regressing genetic distances from the tips to the root of a maximum likelihood (ML) tree against sampling dates (**Fig. 3A, Fig. S7**). We found sufficient temporal structure in our dataset (r^2^=0.51) (**Fig. 3A**). Subsequently, our molecular clock analysis using an exponential parametric tree prior inferred an evolutionary rate of 1.126 ⨯ 10^×3^ (95 % BCI: 1.03–1.23 ⨯ 10^×3^) substitutions per site per year (s/s/y), equating to an average of 33 changes per year. This is within the evolutionary rates estimated for other human coronaviruses (*26-29*). We estimate the date of the common ancestor of the SARS-CoV-2 pandemic to around mid-Nov 2019 (median estimate 17 Nov 2019, 95% BCI: 26 Oct 2019–16 Dec 2019), in line with recent findings (*30, 31*). Similar dates of emergence and rates of evolutionary change were obtained using a more flexible non-parametric coalescent model.

**Fig. 3.**
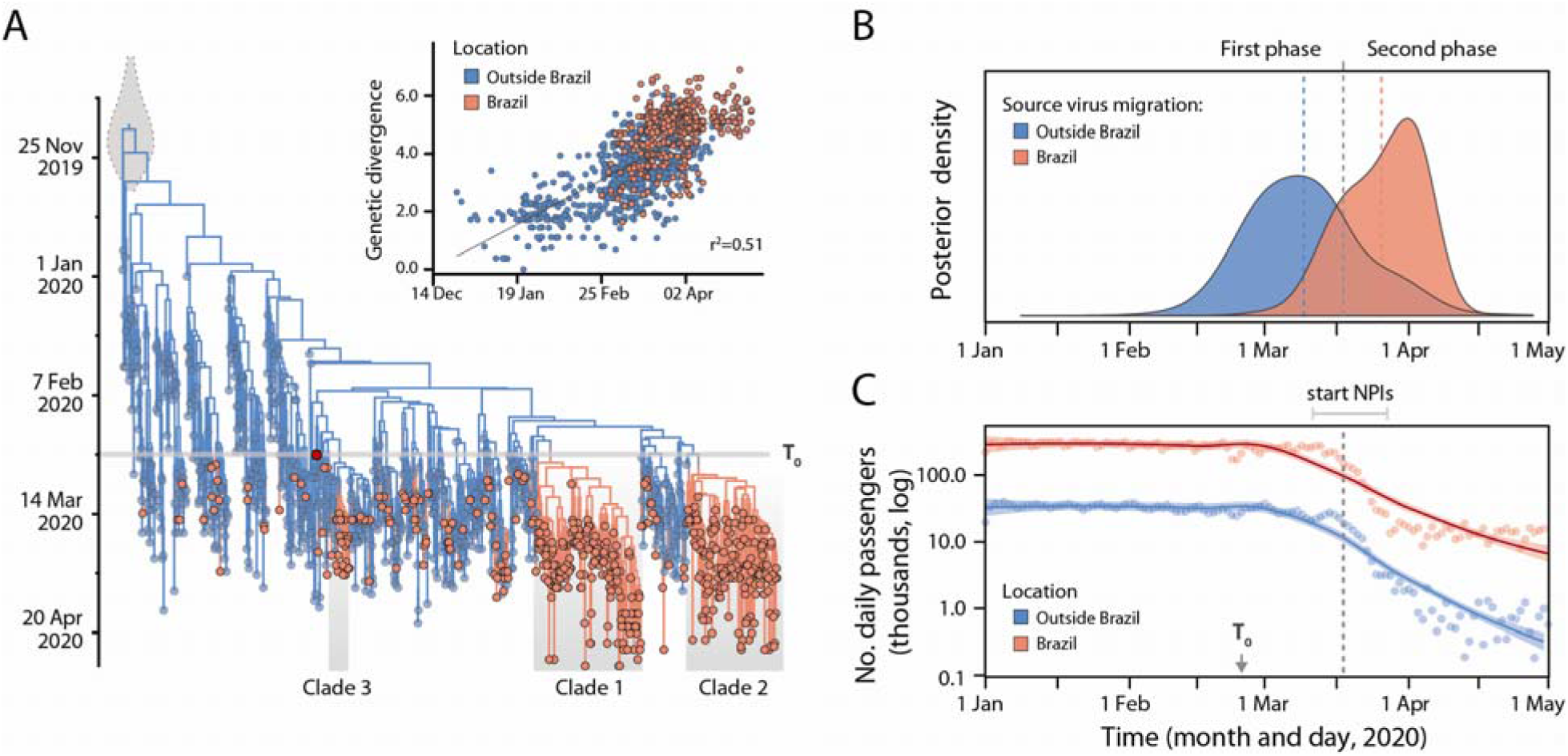
Evolution and spread of SARS-CoV-2 in Brazil. (**A**) Time-resolved maximum clade credibility phylogeny of 1182 SARS-CoV-2 sequences, 490 from Brazil (salmon) and 692 from outside Brazil (blue). The largest Brazilian clades are highlighted in grey (*Clade 1, Clade 2* and *Clade 3*). Detailed dates of emergence for these clades are shown in **Fig. S8**. Inset on the upper left corner shows the root-to-tip regression of genetic divergence against dates of sample collection. (**B**) Dynamics of SARS-CoV-2 import events in Brazil. Dates of international and national (between states) migration events were estimated from sequence data using a phylogeographic approach. The first phase was dominated by virus migrations from outside Brazil while the second phase as marked by virus spread within Brazil. (**C**) Locally estimated scatterplot smoothing (LOESS) of the daily number of international (blue) and national (salmon) air passengers in Brazil in 2020. A detailed annotated phylogeny can be found in **Fig. S8**. T_0_ = date of first reported case in Brazil (25 February 2020).

Time-measured phylogeographic analyses revealed at least 104 (95% BCI: 101–108) international introductions in Brazil (**Fig. 3A, Figs. S8-S9**). This is likely to represent an underestimate of the real number of introductions as we have sequenced, on average, 1 out of 200 confirmed cases (0.5%). Most of these introductions were directed into internationally well-connected states: São Paulo (36% of all imports), Minas Gerais (24%), Ceará (10%) and Rio de Janeiro (8%) (**Fig. S9**). The majority of the Brazilian genomes (75%, *n*=366/490) fall into three monophyletic clades hereafter named as *Clade 1* (*n*=186/490, 38% of all strains from Brazil), *Clade 2* (*n*=161, 33%) and *Clade 3* (*n*=19/490, 4%) (**Fig. 3A, Fig. S10**), which were also identified in a phylogenetic analysis using 13,307 global genomes. We estimate that these clades were introduced from Europe and emerged in Brazil between 22 and 27 February 2020, with upper credible intervals varying from 29 February to 3 March (see also **Fig. S11**). Similar to what has been recently shown in New York City (*32*), this indicates that community-driven transmission became established in Brazil around late February and early March, highlighting that international travel restrictions initiated after this period would have had limited efficacy.

*Clade 1* is characterized by an amino acid mutation in the spike protein (G25088T, reference GenBank accession number, NC_045512.2) and circulates predominantly in São Paulo state (n=159, 85.4%, **Figs. S10 and S12**). *Clade 2* is defined by two amino acid mutations in the ORF6 (T27299C) and nucleoprotein (T29148C); this is the most spatially widespread lineage, with sequences from a total of 16 states in Brazil. *Clade 3* (*n*=19, 4%), is concentrated in Ceará state (*n*=16, 84%) and falls in a global cluster with sequences mainly from Europe. In the Amazon region, where the epidemic is expanding rapidly (*22*), we find evidence for multiple national and international introductions, with 37% (*n*=7/19) of all sequences from Pará and the Amazonas states clustering in *Clade 1* and 32% (*n*=6/19) in *Clade 2*. An interactive visualization of the temporal, geographic and mutational patterns in our data can be found in https://microreact.org/project/eyBufSP8UwVNQqBDjxLEuj.

Although metadata on patient’s travel history is critical to differentiate international importation and local transmission, such data is only available for the first month of the epidemic in Brazil (*15*). We develop an analysis of genetic data to assess the contribution of international vs. national virus lineage movement events through time (**Fig. 3B**). In the first phase of the epidemic, we find an increasing number of international introductions until the 10 Mar 2020 (**Fig. 2B**). According to line-list epidemiological data on the first 1,468 cases in Brazil (*15*), early cases were predominantly acquired from Italy (26%, *n*=70 of 266 unambiguously identified country of infection) and the USA (28%, *n*=76 of 266) during this period). Our analysis suggests that São Paulo, Minas Gerais, Ceára and Rio de Janeiro states, where the three main clades circulate, received the highest proportion of international imports (**Fig. S9**). After this, the estimated number of international imports decreased concomitantly with the decline in the number of international passengers travelling to Brazil (**Figs. 3B and 3C**, see also **Fig. S13**). In contrast, despite the declines in the number of passengers travelling on national flights (**Fig. 3C**), the number of virus lineage movement events within Brazil continued to increase until around 24 March 2020.

To better understand the contribution of virus spread across spatiotemporal scales in Brazil, we use a continuous phylogeographic model that maps phylogenetic nodes to their inferred origin locations (*33*) (**Fig. 4**). We distinguish branches that remain within a state versus those that cross a state to infer the proportion of within and between state measured virus movement.

**Fig. 4.**
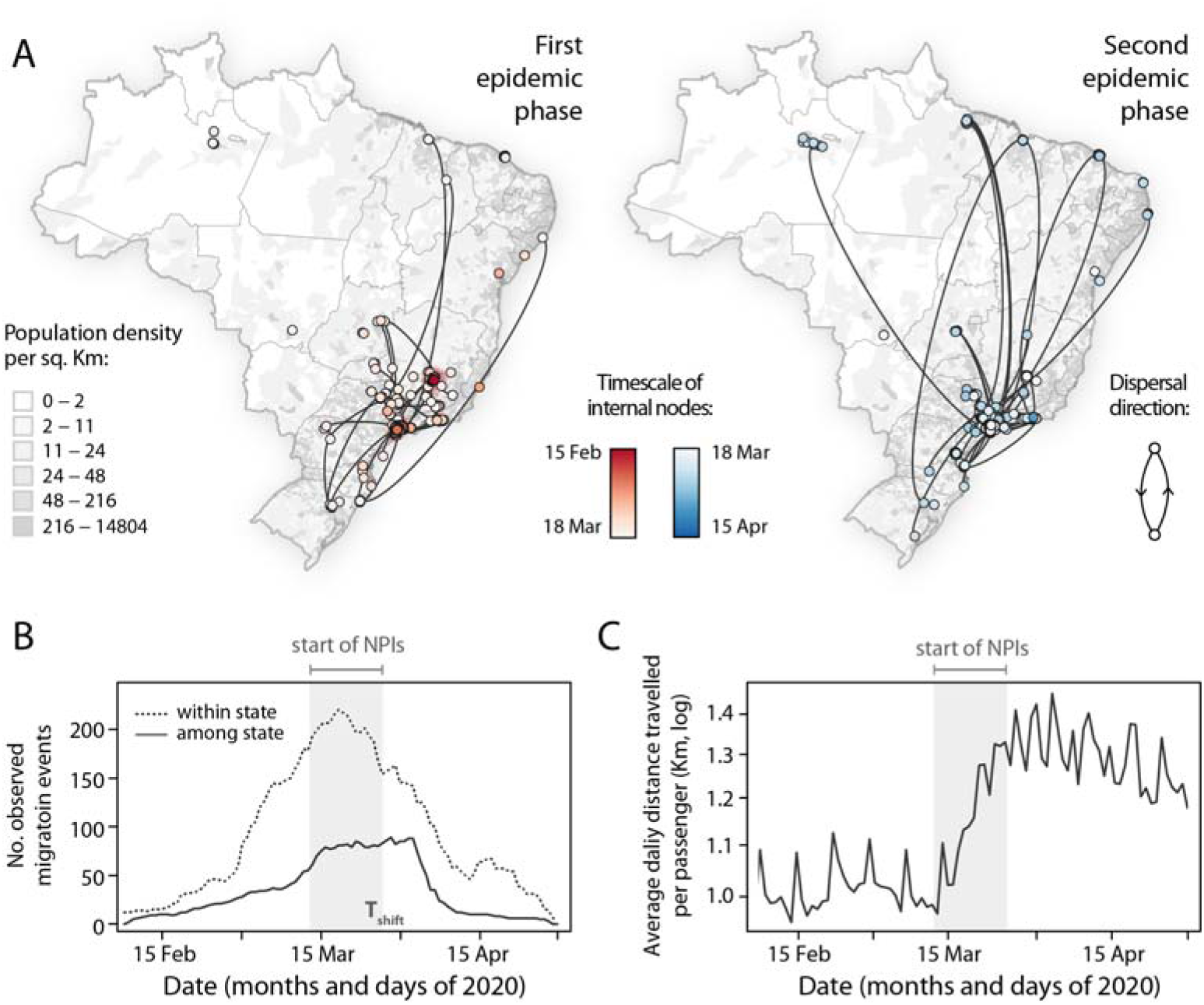
Spread of SARS-CoV-2 in Brazil. (**A**) Spatiotemporal reconstruction of the spread of Brazilian SARS-CoV-2 clusters containing 3 or more sequences during the first phase (left) and the second epidemic phase (right). Circles represent nodes of the MCC phylogeny and are coloured according to their inferred time of occurrence. Shaded areas represent the 80% high posterior density (HPD) interval and depict the uncertainty of the phylogeographic estimates for each node. Solid curved lines denote the links between sequences and the directionality of movement. Sequences belonging to clusters with less than 3 sequences were also plotted in the map with no lines connecting them. Background population density in 2020 for each municipality was obtained from the Brazilian Institute of Geography (https://www.ibge.gov.br/). **Fig. S14** shows a zoomed version of virus spread in the Southeast region. (**B**) Estimated number of within state and between state virus migrations over time. (**C**) Average distance travelled by an air passenger per day in Brazil calculated using openly available data from the National Civil Aviation Agency of Brazil (www.anac.gov.br/en). Light grey boxes indicate starting dates of NPIs across Brazil.

We estimate that during the first epidemic phase, SARS-CoV-2 spread mostly locally and within-state borders. In contrast, the second phase was characterized by long-distance movement events and the ignition of the epidemic outside the southeast region (**Fig. 4A**). Throughout the epidemic, we find that within-state virus movement was on average 2.7-fold more frequent than between-state virus movement. Moreover, our data suggests that within-state virus spread, and to a lesser extent, between-state virus spread, decreased after the implementation of NPIs (**Fig. 4B**). Interestingly, we find that the average route length travelled by passenger increased by 25% during the second phase of the epidemic (**Fig. 4C**), despite a concomitant reduction in the number of passengers flying within Brazil (**Fig. 3C**). These findings emphasize the role of within and between-state mobility as a key driver of both local and inter-regional virus spread, with highly populated and well-connected urban conurbations in southeast region acting as main sources of virus exports within the country (**Fig. S9**).

In conclusion, our study provides the first comprehensive analysis of SARS-CoV-2 spread in Brazil. We found no evidence of community transmission before the first reported case in Brazil, and that large highly connected urban centres drove virus spread across the country. We show that NPIs significantly reduced both virus transmission and spread, but that additional measures are required to maintain transmission at low levels. With recent relaxation of NPIs in Brazil, continued surveillance is required to monitor trends in transmission and the emergence of new lineages in Brazil. Our analysis shows how changes in mobility may impact global and local transmission of SARS-CoV-2, and demonstrates how combining genomic and mobility data can guide traditional surveillance approaches.

## Data Availability

All data, code, and materials used in the analysis are available on our Dryad and GitHub repository (upon acceptance).

## Acknowledgments

A full list acknowledging those involved in the diagnostic and generation of new sequences as part of the CADDE-Genomic-Network can be found in the Supplementary Material. We thank GISAID database for supporting rapid and transparent sharing of genomic data during the COVID-19 pandemic. A full list acknowledging the authors submitting data used in this study can be found in **Data S2**. We thank Lucy Matkin and Josh Quick for logistic support to the CADDE project.

## Supplementary Materials

Materials and Methods

Figs. S1 to S14

Tables S1 to S3

Captions for Data S1 and Data S2

List of Members of the CADDE-Genomic-Network

Full Reference List

